# SARS-CoV-2 spike protein gene variants with N501T and G142D mutation dominated infections in minks in the US

**DOI:** 10.1101/2021.03.18.21253734

**Authors:** Hugh Y. Cai, Allison Cai

## Abstract

Large number of minks were infected with SARS-CoV-2 virus containing the spike protein Y453F mutation in Europe, causing zoonosis concerns. To evaluate the genetic characteristics of the US and Canadian mink-derived SARS-CoV-2 sequences, we analyzed all animal-derived (977), all Canadian (19,529) and US (173,277) SARS-CoV-2 sequences deposited in GISAID from December 2019 to March 12, 2021, and identified 2 dominant novel variants, the N501T-G142D variant and N501T-G142D-F486L variant, in the US mink-derived SARS-CoV-2 sequences. These variants were not found in minks from Canada or other countries. The Y453F mutation was not identified in the mink-derive sequences in the US and Canada. The N501T mutation occurred two months earlier in the human than in the minks in the US, and the novel N501T-G142D variant and N501T-G142D-F486L variant were found in human prior to minks. The result of this study indicates that the novel variants may have evolved during human infection and then transmitted to mink populations in the US.

---

Genomic surveillance of circulating SARS-CoV-2 variants is critical for epidemiologic tracking and infection control. Rapid global spread of a variant harbouring a mutation located on the SARS-CoV-2 spike protein (S protein) has been reported.^6,7^ In the GISAID database of SARS-CoV-2 genomes as of March 17, 2021, 95.7% (758,684 of 792,388) of the SARS-CoV-2 genome has the D614G mutation in the spike protein gene; and 22% (178,186 of 792.388) had N501Y mutation which occurred in the more virulent UK variant (lineage B.1.1.7) with increased transmissibility by 50% and is potentially more virulent;^1^ a similar phenomenon was observed with the South African variant lineage B.1.351^8^ and the Brazil variant lineage B.1.525,^4,9^ both have the N501Y mutation. Large numbers of mutants with Y453F mutations were found in infected mink in Denmark, which might have been spread from human to animal and back to human.^5^ We analyzed the sequences of the SARS-CoV-2 S protein gene collected from mink in the USA and Canada, and identified 2 dominant novel variants with 2 and 3 mutations each.

From the GISAID (Global Initiative on Sharing All Influenza Data, https://www.gisaid.org), we downloaded the following SARS-CoV-2 sequences collected from December 2019 to the date of writing (March 12, 2021): all animal-derived sequences (977) including those from *Canis lupus familiaris, Chlorocebus sabaeus, Felis catus, Manis javanica, Manis pentadactyla, Mus musculus, Mustela lutreola, Neovison vison, Panthera leo, Panthera tigris jacksoni, Rhinolophus affinis, Rhinolophus malayanus and Rhinolophus shameli*; all sequences collected in Canada (19,529) and the US (173,277). The SARS-CoV-2 isolate Wuhan-Hu-1 collected in December 19, 2019 (GenBank accession NC045512) was used as a reference for mutation analysis. All nucleotide position labeling in our study was based on alignment with this sequence. SARS-CoV-2 full genome sequences were multiple aligned with the bioinformatic software, Geneious v.11 (Auckland, New Zealand) using “Geneious Multiple Alignment” and “Map to a Reference Assembly” function. The aligned sequences were visually examined to confirm that they were aligned properly. The variants/SNP were identified by the software automatically and verified by visual confirmation. Short fragments (30 nt) containing the novel mutations identified in our study were used as queries to blast search the local databases consisting of the downloaded sequences to determine the incidence of the novel mutations using the “BLAST” function of the Geneious software. The global incidence of “N501Y” was determined using the “search” function with substitution “N501Y” in GISAID. The global incidence of “N501T” was not determined since the query of substitution “N501T” was not available in GISAID.

The N501Y mutation that was common to UK, South African, and Brazil variants, and the Y453F mutation that widely prevailed in COVID-19 positive mink in Denmark and the Netherlands were absent in all the US and Canadian mink SARS-CoV-2 sequences examined. Interestingly, an N501T mutation was found in almost all US mink SARS-CoV-2 sequences (100 of 101; 99%); furthermore, all N501T mutant sequences, except 2 of poor quality, had the G142D mutation (Supplementary Table 1). Based on GISAID records, all US mink SARS-CoV-2 sequences were collected from Michigan and Wisconsin in October, 2020. We found that, except 1 sequences with poor quality, all (54) Michigan mink sequences had the F486L mutation, and all (55) Michigan mink sequences had the Y144 deletion, in addition to the N501T and G142D mutations (Supplementary Table 1). Hereafter, we designated the sequences with N501T and G142D, but without F486L, mutations as the N501T-G142D variant (WI mink variant); and the ones with all 3 novel mutations as the N501T-G142D-F486L variant (MI mink variant). In addition to the above 3 mutations and 1 deletion in Spike protein, G614D, the mutation that dominated the SARS-CoV-2 sequences, existed in all US and Canadian mink-derived sequences (Supplementary Table 1, Supplementary Table 2). No N501T-G142D nor N501T-G142D-F486L variants were found in mink from Canada or other countries.

Outside the US, among 335 Netherlands mink-derived SARS-CoV-2 S protein gene sequences, 5 collected from April to June 2020 had the N501T mutation without G142D or F486L mutations (Supplementary Table 3), which is consistent with a previous report on N501T on Netherlands mink-derived sequences.^2^ We also found F486L mutation in 60% (204 of 335) Netherlands mink-derived SARS-CoV-2 S protein gene sequences, none of which with N501T-G142D variants. The F486L mutation was unique to Netherlands mink-derived sequences causing by nucleotide mutation T23018C, different from the F486L mutation found in the US mink-derived sequences due to nucleotide T23020G mutation. Among 454 Denmark mink SARS-CoV-2 sequences, 3 had N501T mutations as described earlier, none had G142D or F486L mutations (Supplementary Table 3).

In order to determine the origin of the 2 novel variants, we analyzed all human-derived SARS-CoV-2 sequences collected from the US and Canada. Three WI N501T-G142D variant and 2 MI N501T-G142D-F486L variant sequences were identified from the US human-derived sequences collected from Wisconsin on October 3, 2020 and Michigan on October 6, 2020, and they had all the mutations as those described above for the mink sequences except lacking of the Y144 deletion in S gene. Furthermore, we identified N501T mutants without G142D and F486L in US human-derived sequences collected as early as August 2020, which was earlier than the mink-derived sequences collected in October, 2020 (Supplementary Table 4, Fig. 1). In Canada, the N501T mutation only existed in human-derived sequences (Supplementary Table 2). The N501T mutation was described in 2 Italian human-derived sequences collected in August 2020.^3^ We therefore speculate that the N501T mutation may have evolved during human infection and was then transmitted to mink populations in the US, and it is less likely that the mutations occurred in mink population after infected with human strains of the virus. The fact that the two novel variants were found in almost all infected minks in the US in short period of times within days also support this speculation.

**Fig. 1.**
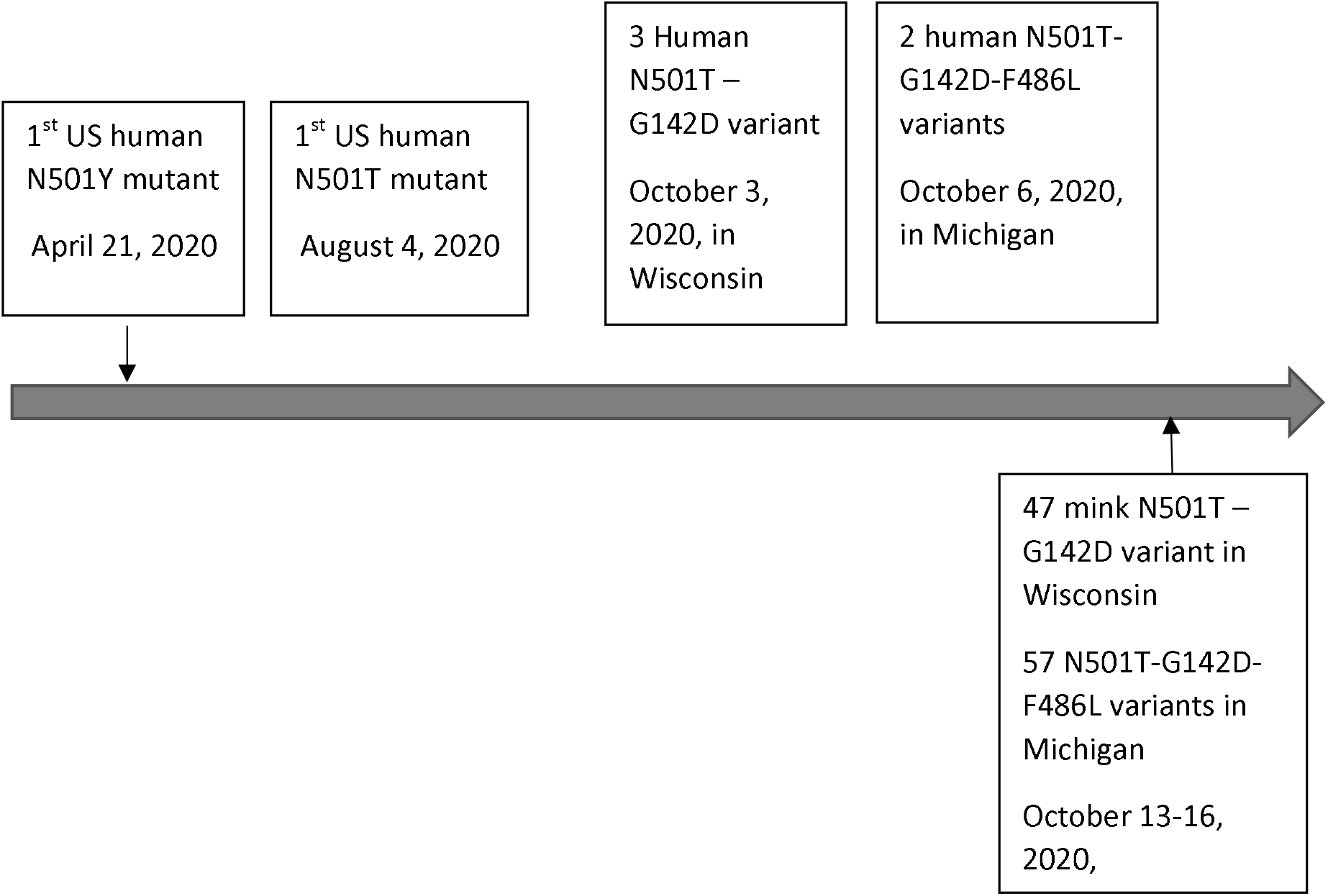
Emerging timeline of human-derived (above the timeline) and mink-derived (below the timeline) SARS-CoV-2 spike protein N501T-G142D and N501T-G142D-F486L variants in the US. Timeline scale is not in proportion.

To evaluate the transmissibility of N501T mutants, the incidence of N501T and N501Y mutants was enumerated from US and Canadian GISAID SARS-CoV-2 sequences collected up to March 12, 2021: 2.4% (4,097 of 173,277) and 0.77% (1,339 of 173,277) of US sequences had the N501Y and N501T maturation, respectively; and 1.6% (315/19,529) and 0.06% (12/19,529) of Canadian sequences had the N501Y and the N501T mutation, respectively (Supplementary Table 5). Comparatively, the incidence of N501Y is much lower in the US and Canada than globally. Our analysis results indicate that the N501T mutants had weaker transmissibility compared to the N501Y mutants that include UK B.1.17, South Africa B.1.351, and Brazil P.1. However, it cannot be ruled out that the different time of introduction of N501Y and N501T (April 2020 vs August 2020) may have affected the occurrences of the 2 mutants (Fig. 1).

It has been described that the mutations at S protein N501 can significantly increase the transmissibility, and likely virulence of SARS-CoV-2; the UK variant N501Y is an example. It was predicted that a single N501T mutation might significantly enhance the binding affinity between 2019-nCoV RBD and human ACE2.^10^ Our analysis revealed that the N501T and G142DL mutations occurred in 99% of mink-derived sequences collected in the US; all, except one with poor quality, mink sequences collected from Michigan also contained the F486L mutation. The large number of new variants occurring in the mink population warrants further study on how these changes affected their interaction with the ACE2 receptor, and thereby the transmissibility, virulence, and immunogenicity in humans and mink. It is important to monitor the emerging new variants and determine their impact on human and animal health.

## Supporting information

supplementary acknowledgement

## Data Availability

All sequence data used in this study were available from the GISAID hCov-19 Database. GISAID sequences with N501T, G142D and F486L mutations are listed in Supplementary Tables.

https://www.gisaid.org/

## Acknowledgement

We gratefully acknowledge the authors, originating and submitting laboratories of the sequences from GISAID’s hCov-19 Database on which this research is based. See supplementary acknowledgement for details.

## Declaration of conflicting interests

The authors declared no potential conflicts of interest with respect to the research, authorship, and/or publication of this article.

## Funding Support

The authors received no financial support for the research, authorship, and/or publication of this article.

## Supplementary material

Supplementary material for this article is available online.

## Notes

### Competing Interest Statement

The authors have declared no competing interest.

### Clinical Trial

The study was performed on SARS-CoV-2 genome sequences deposited in public database (GISAID). It is not a clinical trial.

### Author Declarations

This study was performed on publicly available SARS-CoV-2 genome sequences only.

### Summary of Updates

Added information on Denmark and Netherlands and Denmark mink-derived SARS-CoV-2 sequences (Line 78-94).

