## Supplementary Table 1 for "SARS-CoV-2 spike protein gene variants with N501T and G142D mutation dominated infections in minks in the US"

Supplementary Table 1: SARS-Cov-2 Spike protein mutation N501T and G142D variants in US minks

| GISAID Sequences | Collection date | Location | Mutations in S gene | | | |
|---|---|---|---|---|---|---|
| | | | G142 | F486 | N501 | D614 |
| hCoV-19/mink/USA/MI-CDC-IHO1-6827/2020\|EPI_ISL_925341\|2020-10-13 | 10/13/2020 | Michigan | G142D | F486L | N501T | D614G |
| hCoV-19/mink/USA/MI-CDC-IHOZ-6866/2020\|EPI_ISL_925354\|2020-10-13 | 10/13/2020 | Michigan | G142D | F486L | N501T | D614G |
| hCoV-19/mink/USA/MI-CDC-IHP1-6869/2020\|EPI_ISL_925342\|2020-10-13 | 10/13/2020 | Michigan | G142D | F486L | N501T | D614G |
| hCoV-19/mink/USA/MI-CDC-IHP2-6870/2020\|EPI_ISL_925353\|2020-10-13 | 10/13/2020 | Michigan | G142D | F486L | N501T | D614G |
| hCoV-19/mink/USA/MI-CDC-IHPJ-6890/2020\|EPI_ISL_925314\|2020-10-13 | 10/13/2020 | Michigan | G142D | F486L | N501T | D614G |
| hCoV-19/mink/USA/MI-CDC-IHQ4-6914/2020\|EPI_ISL_925343\|2020-10-13 | 10/13/2020 | Michigan | G142D | F486L | N501T | D614G |
| hCoV-19/mink/USA/MI-CDC-IHQ7-6918/2020\|EPI_ISL_925344\|2020-10-13 | 10/13/2020 | Michigan | G142D | F486L | N501T | D614G |
| hCoV-19/mink/USA/MI-CDC-IHQE-6926/2020\|EPI_ISL_925355\|2020-10-13 | 10/13/2020 | Michigan | G142D | F486L | N501T | D614G |
| hCoV-19/mink/USA/MI-CDC-IHQV-6946/2020\|EPI_ISL_925336\|2020-10-13 | 10/13/2020 | Michigan | G142D | F486L | N501T | D614G |
| hCoV-19/mink/USA/MI-CDC-IHRD-6976/2020\|EPI_ISL_925345\|2020-10-14 | 10/14/2020 | Michigan | G142D | F486L | N501T | D614G |
| hCoV-19/mink/USA/MI-CDC-IHRM-6977/2020\|EPI_ISL_925356\|2020-10-14 | 10/14/2020 | Michigan | G142D | F486L | N501T | D614G |
| hCoV-19/mink/USA/MI-CDC-IHRU-6987/2020\|EPI_ISL_925315\|2020-10-14 | 10/14/2020 | Michigan | G142D | F486L | N501T | D614G |
| hCoV-19/mink/USA/MI-CDC-IHS4-6998/2020\|EPI_ISL_925357\|2020-10-14 | 10/14/2020 | Michigan | G142D | F486L | N501T | D614G |
| hCoV-19/mink/USA/MI-CDC-IHSI-7015/2020\|EPI_ISL_925346\|2020-10-14 | 10/14/2020 | Michigan | G142D | F486L | N501T | D614G |
| hCoV-19/mink/USA/MI-CDC-IHSO-7022/2020\|EPI_ISL_925347\|2020-10-14 | 10/14/2020 | Michigan | G142D | F486L | N501T | D614G |
| hCoV-19/mink/USA/MI-CDC-IHTR-7067/2020\|EPI_ISL_925348\|2020-10-14 | 10/14/2020 | Michigan | G142D | F486L | N501T | D614G |
| hCoV-19/mink/USA/MI-CDC-IHTS-7068/2020\|EPI_ISL_925358\|2020-10-14 | 10/14/2020 | Michigan | G142D | F486L | N501T | D614G |
| hCoV-19/mink/USA/MI-CDC-IHU3-7081/2020\|EPI_ISL_925349\|2020-10-14 | 10/14/2020 | Michigan | G142D | F486L | N501T | D614G |
| hCoV-19/mink/USA/MI-CDC-IHU4-7082/2020\|EPI_ISL_925334\|2020-10-14 | 10/14/2020 | Michigan | G142D | F486L | N501T | D614G |
| hCoV-19/mink/USA/MI-CDC-IHU5-7083/2020\|EPI_ISL_925360\|2020-10-14 | 10/14/2020 | Michigan | G142D | F486L | N501T | D614G |
| hCoV-19/mink/USA/MI-CDC-IHU6-7085/2020\|EPI_ISL_925337\|2020-10-13 | 10/14/2020 | Michigan | G142D | F486L | N501T | D614G |
| hCoV-19/mink/USA/MI-CDC-IHU7-7086/2020\|EPI_ISL_925338\|2020-10-14 | 10/14/2020 | Michigan | G142D | F486L | N501T | D614G |
| hCoV-19/mink/USA/MI-CDC-IHUD-7093/2020\|EPI_ISL_925350\|2020-10-14 | 10/14/2020 | Michigan | G142D | F486L | N501T | D614G |
| hCoV-19/mink/USA/MI-CDC-IHUE-7094/2020\|EPI_ISL_925335\|2020-10-14 | 10/14/2020 | Michigan | G142D | F486L | N501T | D614G |
| hCoV-19/mink/USA/MI-CDC-IHUH-7098/2020\|EPI_ISL_925339\|2020-10-15 | 10/15/2020 | Michigan | G142D | F486L | N501T | D614G |
| hCoV-19/mink/USA/MI-CDC-IHUI-7099/2020\|EPI_ISL_925340\|2020-10-16 | 10/16/2020 | Michigan | G142D | F486L | N501T | D614G |
| hCoV-19/mink/USA/MI-CDC-IHUK-7101/2020\|EPI_ISL_925351\|2020-10-14 | 10/14/2020 | Michigan | G142D | F486L | N501T | D614G |
| hCoV-19/mink/USA/MI-CDC-IHUL-7102/2020\|EPI_ISL_925359\|2020-10-14 | 10/14/2020 | Michigan | G142D | F486L | N501T | D614G |
| hCoV-19/mink/USA/MI-CDC-IHUR-7108/2020\|EPI_ISL_925352\|2020-10-15 | 10/15/2020 | Michigan | G142D | F486L | N501T | D614G |
| hCoV-19/mink/USA/MI-CDC-IHVC-7129/2020\|EPI_ISL_925318\|2020-10-15 | 10/15/2020 | Michigan | G142D | F486L | N501T | D614G |

| | | | | | | |
|---|---|---|---|---|---|---|
| hCoV-19/mink/USA/MI-CDC-IHVH-7139/2020\|EPI_ISL_925323\|2020-10-15 | 10/15/2020 | Michigan | G142D | F486L | N501T | D614G |
| hCoV-19/mink/USA/MI-CDC-IHVN-7173/2020\|EPI_ISL_925316\|2020-10-15 | 10/15/2020 | Michigan | G142D | F486L | N501T | D614G |
| hCoV-19/mink/USA/MI-CDC-IHVT-7174/2020\|EPI_ISL_925330\|2020-10-15 | 10/15/2020 | Michigan | G142D | F486L | N501T | D614G |
| hCoV-19/mink/USA/MI-CDC-IHVZ-7167/2020\|EPI_ISL_925325\|2020-10-15 | 10/15/2020 | Michigan | G142D | F486L | N501T | D614G |
| hCoV-19/mink/USA/MI-CDC-IHW2-3526/2020\|EPI_ISL_925320\|2020-10-15 | 10/15/2020 | Michigan | G142D | F486L | N501T | D614G |
| hCoV-19/mink/USA/MI-CDC-IHW5-7133/2020\|EPI_ISL_925322\|2020-10-15 | 10/15/2020 | Michigan | G142D | F486L | N501T | D614G |
| hCoV-19/mink/USA/MI-CDC-IHWB-7153/2020\|EPI_ISL_925308\|2020-10-15 | 10/15/2020 | Michigan | G142D | F486L | WT/N501T | D614G |
| hCoV-19/mink/USA/MI-CDC-IHWC-7160/2020\|EPI_ISL_925324\|2020-10-15 | 10/15/2020 | Michigan | G142D | F486L | N501T | D614G |
| hCoV-19/mink/USA/MI-CDC-IHWU-3519/2020\|EPI_ISL_925319\|2020-10-15 | 10/15/2020 | Michigan | G142D | F486L | N501T | D614G |
| hCoV-19/mink/USA/MI-CDC-IHX8-7146/2020\|EPI_ISL_925309\|2020-10-15 | 10/15/2020 | Michigan | G142D | F486L | N501T | D614G |
| hCoV-19/mink/USA/MI-CDC-IHXX-7228/2020\|EPI_ISL_925326\|2020-10-16 | 10/16/2020 | Michigan | G142D | F486L | N501T | D614G |
| hCoV-19/mink/USA/MI-CDC-IHXZ-7239/2020\|EPI_ISL_925333\|2020-10-16 | 10/16/2020 | Michigan | G142D | F486L | N501T | D614G |
| hCoV-19/mink/USA/MI-CDC-IHYE-7249/2020\|EPI_ISL_925317\|2020-10-16 | 10/16/2020 | Michigan | G142D | F486L | N501T | D614G |
| hCoV-19/mink/USA/MI-CDC-IHYF-7184/2020\|EPI_ISL_925331\|2020-10-16 | 10/16/2020 | Michigan | G142D | F486L | N501T | D614G |
| hCoV-19/mink/USA/MI-CDC-IHZF-7215/2020\|EPI_ISL_925332\|2020-10-16 | 10/16/2020 | Michigan | G142D | F486L | N501T | D614G |
| hCoV-19/mink/USA/MI-CDC-IHZZ-7327/2020\|EPI_ISL_925312\|2020-10-16 | 10/16/2020 | Michigan | G142D | F486L | N501T | D614G |
| hCoV-19/mink/USA/MI-CDC-II0C-7328/2020\|EPI_ISL_925310\|2020-10-17 | 10/17/2020 | Michigan | G142D | F486L | N501T | D614G |
| hCoV-19/mink/USA/MI-CDC-II12-7329/2020\|EPI_ISL_925313\|2020-10-18 | 10/16/2020 | Michigan | G142D | F486L | N501T | D614G |
| hCoV-19/mink/USA/MI-CDC-II16-7296/2020\|EPI_ISL_925329\|2020-10-16 | 10/16/2020 | Michigan | G142D | F486L | N501T | D614G |
| hCoV-19/mink/USA/MI-CDC-II19-7314/2020\|EPI_ISL_925311\|2020-10-18 | 10/18/2020 | Michigan | G142D | F486L | N501T | D614G |
| hCoV-19/mink/USA/MI-CDC-II1E-7315/2020\|EPI_ISL_925327\|2020-10-16 | 10/16/2020 | Michigan | G142D | F486L | N501T | D614G |
| hCoV-19/mink/USA/MI-CDC-II1G-3562/2020\|EPI_ISL_925328\|2020-10-16 | 10/16/2020 | Michigan | G142D | F486L | N501T | D614G |
| hCoV-19/mink/USA/MI-CDC-II1O-7265/2020\|EPI_ISL_925307\|2020-10-16 | 10/16/2020 | Michigan | G142D | ND* | WT** | D614G |
| hCoV-19/mink/USA/MI-CDC-II1P-3558/2020\|EPI_ISL_925321\|2020-10-16 | 10/16/2020 | Michigan | G142D | F486L | N501T | D614G |
| hCoV-19/mink/USA/WI-CDC-CWOF-2204/2020\|EPI_ISL_1014965\|2020-10-15 | 10/15/2020 | Wisconsin | G142D | WT | N501T | D614G |
| hCoV-19/mink/USA/WI-CDC-CWOZ-2205/2020\|EPI_ISL_1014950\|2020-10-15 | 10/15/2020 | Wisconsin | G142D | WT | N501T | D614G |
| hCoV-19/mink/USA/WI-CDC-CWPJ-2206/2020\|EPI_ISL_1014943\|2020-10-15 | 10/15/2020 | Wisconsin | G142D | WT | N501T | D614G |
| hCoV-19/mink/USA/WI-CDC-CWQ5-2422/2020\|EPI_ISL_1014974\|2020-10-15 | 10/15/2020 | Wisconsin | G142D | WT | N501T | D614G |
| hCoV-19/mink/USA/WI-CDC-CWR7-2209/2020\|EPI_ISL_1014951\|2020-10-15 | 10/15/2020 | Wisconsin | G142D | WT | N501T | D614G |
| hCoV-19/mink/USA/WI-CDC-CWTH-2425/2020\|EPI_ISL_1014946\|2020-10-15 | 10/15/2020 | Wisconsin | G142D | WT | N501T | D614G |
| hCoV-19/mink/USA/WI-CDC-CWV3-2216/2020\|EPI_ISL_1014952\|2020-10-15 | 10/15/2020 | Wisconsin | G142D | WT | N501T | D614G |
| hCoV-19/mink/USA/WI-CDC-CWWU-2430/2020\|EPI_ISL_1014945\|2020-10-15 | 10/15/2020 | Wisconsin | ND | WT | N501T | D614G |
| hCoV-19/mink/USA/WI-CDC-CWXY-2431/2020\|EPI_ISL_1014975\|2020-10-15 | 10/15/2020 | Wisconsin | G142D | WT | N501T | D614G |
| hCoV-19/mink/USA/WI-CDC-CWYZ-2306/2020\|EPI_ISL_1014955\|2020-10-15 | 10/15/2020 | Wisconsin | G142D | WT | N501T | D614G |

| | | | | | | |
|---|---|---|---|---|---|---|
| hCoV-19/mink/USA/WI-CDC-CX0O-2226/2020\|EPI_ISL_1014953\|2020-10-15 | 10/15/2020 | Wisconsin | G142D | WT | N501T | D614G |
| hCoV-19/mink/USA/WI-CDC-CX2X-2436/2020\|EPI_ISL_1014948\|2020-10-15 | 10/15/2020 | Wisconsin | ND | WT | N501T | D614G |
| hCoV-19/mink/USA/WI-CDC-CX4L-2439/2020\|EPI_ISL_1014941\|2020-10-15 | 10/15/2020 | Wisconsin | G142D | WT | N501T | D614G |
| hCoV-19/mink/USA/WI-CDC-CX5N-2235/2020\|EPI_ISL_1014966\|2020-10-15 | 10/15/2020 | Wisconsin | G142D | WT | N501T | D614G |
| hCoV-19/mink/USA/WI-CDC-CX6R-2237/2020\|EPI_ISL_1014947\|2020-10-15 | 10/15/2020 | Wisconsin | G142D | WT | N501T | D614G |
| hCoV-19/mink/USA/WI-CDC-CX7Y-2442/2020\|EPI_ISL_1014956\|2020-10-15 | 10/15/2020 | Wisconsin | G142D | WT | N501T | D614G |
| hCoV-19/mink/USA/WI-CDC-CX8H-2443/2020\|EPI_ISL_1014957\|2020-10-15 | 10/15/2020 | Wisconsin | G142D | WT | N501T | D614G |
| hCoV-19/mink/USA/WI-CDC-CX9K-2242/2020\|EPI_ISL_1014954\|2020-10-15 | 10/15/2020 | Wisconsin | G142D | WT | N501T | D614G |
| hCoV-19/mink/USA/WI-CDC-CXA4-2243/2020\|EPI_ISL_1014944\|2020-10-15 | 10/15/2020 | Wisconsin | G142D | WT | N501T | D614G |
| hCoV-19/mink/USA/WI-CDC-CXAN-2244/2020\|EPI_ISL_1014967\|2020-10-15 | 10/15/2020 | Wisconsin | G142D | WT | N501T | D614G |
| hCoV-19/mink/USA/WI-CDC-CXB7-2245/2020\|EPI_ISL_1014942\|2020-10-15 | 10/15/2020 | Wisconsin | G142D | WT | N501T | D614G |
| hCoV-19/mink/USA/WI-CDC-CXHY-2454/2020\|EPI_ISL_1014958\|2020-10-15 | 10/15/2020 | Wisconsin | G142D | WT | N501T | D614G |
| hCoV-19/mink/USA/WI-CDC-CXP3-2270/2020\|EPI_ISL_1014968\|2020-10-14 | 10/14/2020 | Wisconsin | G142D | WT | N501T | D614G |
| hCoV-19/mink/USA/WI-CDC-CXPO-2271/2020\|EPI_ISL_1014969\|2020-10-14 | 10/14/2020 | Wisconsin | G142D | WT | N501T | D614G |
| hCoV-19/mink/USA/WI-CDC-CXQA-2462/2020\|EPI_ISL_1014959\|2020-10-14 | 10/14/2020 | Wisconsin | G142D | WT | N501T | D614G |
| hCoV-19/mink/USA/WI-CDC-CXQU-2463/2020\|EPI_ISL_1014976\|2020-10-13 | 10/13/2020 | Wisconsin | G142D | WT | N501T | D614G |
| hCoV-19/mink/USA/WI-CDC-CXRE-2464/2020\|EPI_ISL_1014960\|2020-10-13 | 10/13/2020 | Wisconsin | G142D | WT | N501T | D614G |
| hCoV-19/mink/USA/WI-CDC-CXRY-2465/2020\|EPI_ISL_1014977\|2020-10-13 | 10/13/2020 | Wisconsin | G142D | WT | N501T | D614G |
| hCoV-19/mink/USA/WI-CDC-CXSH-2466/2020\|EPI_ISL_1014961\|2020-10-13 | 10/13/2020 | Wisconsin | G142D | WT | N501T | D614G |
| hCoV-19/mink/USA/WI-CDC-CXT2-2467/2020\|EPI_ISL_1014978\|2020-10-13 | 10/13/2020 | Wisconsin | G142D | WT | N501T | D614G |
| hCoV-19/mink/USA/WI-CDC-CXTJ-2278/2020\|EPI_ISL_1014970\|2020-10-13 | 10/13/2020 | Wisconsin | G142D | WT | N501T | D614G |
| hCoV-19/mink/USA/WI-CDC-CXU6-2469/2020\|EPI_ISL_1014979\|2020-10-13 | 10/13/2020 | Wisconsin | G142D | WT | N501T | D614G |
| hCoV-19/mink/USA/WI-CDC-CXUQ-2470/2020\|EPI_ISL_1014962\|2020-10-13 | 10/13/2020 | Wisconsin | G142D | WT | N501T | D614G |
| hCoV-19/mink/USA/WI-CDC-CXVA-2471/2020\|EPI_ISL_1014980\|2020-10-13 | 10/13/2020 | Wisconsin | G142D | WT | N501T | D614G |
| hCoV-19/mink/USA/WI-CDC-CXVU-2472/2020\|EPI_ISL_1014981\|2020-10-13 | 10/13/2020 | Wisconsin | G142D | WT | N501T | D614G |
| hCoV-19/mink/USA/WI-CDC-CXWE-2473/2020\|EPI_ISL_1014982\|2020-10-13 | 10/13/2020 | Wisconsin | G142D | WT | N501T | D614G |
| hCoV-19/mink/USA/WI-CDC-CXWY-2474/2020\|EPI_ISL_1014963\|2020-10-13 | 10/13/2020 | Wisconsin | G142D | WT | N501T | D614G |
| hCoV-19/mink/USA/WI-CDC-CXXI-2475/2020\|EPI_ISL_1014983\|2020-10-13 | 10/13/2020 | Wisconsin | G142D | WT | N501T | D614G |
| hCoV-19/mink/USA/WI-CDC-CXY2-2476/2020\|EPI_ISL_1014984\|2020-10-13 | 10/13/2020 | Wisconsin | G142D | WT | N501T | D614G |
| hCoV-19/mink/USA/WI-CDC-CXYL-2477/2020\|EPI_ISL_1014985\|2020-10-13 | 10/13/2020 | Wisconsin | G142D | WT | N501T | D614G |
| hCoV-19/mink/USA/WI-CDC-CXZ6-2478/2020\|EPI_ISL_1014986\|2020-10-13 | 10/13/2020 | Wisconsin | G142D | WT | N501T | D614G |
| hCoV-19/mink/USA/WI-CDC-CXZO-2289/2020\|EPI_ISL_1014971\|2020-10-13 | 10/13/2020 | Wisconsin | G142D/G142Y | WT | N501T | D614G |
| hCoV-19/mink/USA/WI-CDC-CY09-2480/2020\|EPI_ISL_1014987\|2020-10-13 | 10/13/2020 | Wisconsin | G142D | WT | N501T | D614G |
| hCoV-19/mink/USA/WI-CDC-CY0S-2291/2020\|EPI_ISL_1014972\|2020-10-13 | 10/13/2020 | Wisconsin | G142D | WT | N501T | D614G |

| | | | | | | |
|---|---|---|---|---|---|---|
| hCoV-19/mink/USA/WI-CDC-CY1D-2482/2020\|EPI_ISL_1014949\|2020-10-13 | 10/13/2020 | Wisconsin | G142D | WT | N501T | D614G |
| hCoV-19/mink/USA/WI-CDC-CY1V-2293/2020\|EPI_ISL_1014964\|2020-10-13 | 10/13/2020 | Wisconsin | G142D | WT | N501T | D614G |
| hCoV-19/mink/USA/WI-CDC-CY2G-2294/2020\|EPI_ISL_1014973\|2020-10-13 | 10/13/2020 | Wisconsin | G142D | WT | N501T | D614G |

*ND: not determined due to poor sequence quality in the region

**WT:  no mutation compared to NC_045512.2 Severe acute respiratory syndrome coronavirus 2 isolate Wuhan-Hu-1, complete genome
