## Supplementary Table 2 for "SARS-CoV-2 spike protein gene variants with N501T and G142D mutation dominated infections in minks in the US"

Supplementary Table 2. SARS-CoV-2 Spike protein N501T varaints in Canadian sequences

| | Host | Collection date | Mutation in S gene | | | |
|---|---|---|---|---|---|---|
| | | | G142 | F486 | N501 | D614 |
| hCoV-19/mink/Canada/D/2020\|EPI_ISL_717717\|2020-12-04 | Mink | 2020-12 | WT* | WT | WT | D614G |
| hCoV-19/mink/Canada/C/2020\|EPI_ISL_717716\|2020-12-04 | Mink | 2020-12 | WT | WT | WT | D614G |
| hCoV-19/mink/Canada/B/2020\|EPI_ISL_717715\|2020-12-04 | Mink | 2020-12 | WT | WT | WT | D614G |
| hCoV-19/mink/Canada/A/2020\|EPI_ISL_717714\|2020-12-04 | Mink | 2020-12 | WT | F486L** | WT | D614G |
| hCoV-19/Canada/ON-UHTC_0390/2020\|EPI_ISL_698100\|2020-11-24 | Human | 2020-11 | WT | WT | N501T | D614G |
| hCoV-19/Canada/ON-PHL-21-00780/2021\|EPI_ISL_966110\|2021-01 | Human | 2021-01 | WT | WT | N501T | D614G |
| hCoV-19/Canada/ON-PHL-20-03664/2020\|EPI_ISL_775102\|2020-11 | Human | 2020-11 | WT | WT | N501T | D614G |
| hCoV-19/Canada/ON-PHL-20-03660/2020\|EPI_ISL_775098\|2020-11 | Human | 2020-11 | WT | WT | N501T | D614G |
| hCoV-19/Canada/NS-NML-5535/2020\|EPI_ISL_1055694\|2020-12-22 | Human | 2020-12 | WT | WT | N501T | D614G |
| hCoV-19/Canada/NS-NML-5251/2020\|EPI_ISL_1055536\|2020-12-26 | Human | 2020-12 | WT | WT | N501T | D614G |
| hCoV-19/Canada/NS-NML-5226/2020\|EPI_ISL_1055517\|2020-12-12 | Human | 2020-12 | WT | WT | N501T | D614G |
| hCoV-19/Canada/NS-NML-5219/2020\|EPI_ISL_1055511\|2020-12-14 | Human | 2020-12 | WT | WT | N501T | D614G |
| hCoV-19/Canada/NS-NML-5217/2020\|EPI_ISL_1055509\|2020-12-15 | Human | 2020-12 | WT | WT | N501T | D614G |
| hCoV-19/Canada/NS-NML-5216/2020\|EPI_ISL_1055508\|2020-12-15 | Human | 2020-12 | WT | WT | N501T | D614G |
| hCoV-19/Canada/NS-NML-5215/2020\|EPI_ISL_1055507\|2020-12-15 | Human | 2020-12 | WT | WT | N501T | D614G |
| hCoV-19/Canada/NS-NML-5195/2020\|EPI_ISL_1055491\|2020-12-21 | Human | 2020-12 | WT | WT | N501T | D614G |

*WT:  no mutation compared to NC_045512.2 Severe acute respiratory syndrome coronavirus 2 isolate Wuhan-Hu-1, complete genome

** F486L mutatons caused necleotide T23018C
