## Supplementary Table 3 for "SARS-CoV-2 spike protein gene variants with N501T and G142D mutation dominated infections in minks in the US"

Supplementary Table 3: SARS-Cov-2 Spike protein mutation N501T in Denmark and Netherlands mink-derived sequences

| GISAID Sequences | Collection date | Mutations in S gene | | | |
|---|---|---|---|---|---|
| | | G142 | F486 | N501 | D614 |
| hCoV-19/mink/Netherlands/NB-EMC-8-16/2020\|EPI_ISL_523115\|2020-06-01 | 6/1/2020 | WT* | WT | N501T | D614G |
| hCoV-19/mink/Netherlands/NB-EMC-6-14/2020\|EPI_ISL_523099\|2020-05-30 | 5/30/2020 | WT | WT | N501T | D614G |
| hCoV-19/mink/Netherlands/NB-EMC-5-13/2020\|EPI_ISL_523097\|2020-05-30 | 5/30/2020 | WT | WT | N501T | WT |
| hCoV-19/mink/Netherlands/NB-EMC-1-2/2020\|EPI_ISL_523004\|2020-04-28 | 4/28/2020 | WT | WT | N501T | D614G |
| hCoV-19/mink/Netherlands/NB01_02KS/2020\|EPI_ISL_447624\|2020-04-29 | 4/29/2020 | WT | WT | N501T | D614G |
| hCoV-19/mink/Denmark/mDK-174/2020\|EPI_ISL_683025\|2020-11-04 | 11/4/2020 | WT | WT | N501T | D614G |
| hCoV-19/mink/Denmark/mDK-173/2020\|EPI_ISL_683024\|2020-11-04 | 11/4/2020 | WT | WT | N501T | D614G |
| hCoV-19/mink/Denmark/mDK-172/2020\|EPI_ISL_683023\|2020-11-04 | 11/4/2020 | WT | WT | N501T | D614G |

*WT: no mutation compared to NC_045512.2 Severe acute respiratory syndrome coronavirus 2 isolate Wuhan-Hu-1, complete genome
