## Supplementary Table 4 for "SARS-CoV-2 spike protein gene variants with N501T and G142D mutation dominated infections in minks in the US"

Supplementary Table 4: Earlist SARS-CoV-2 spike protein  N501T varaints in US human infection

| | Collection date | Location | Mutation in S gene | | | |
|---|---|---|---|---|---|---|
| | | | G142 | F486 | N501 | D614G |
| hCoV-19/USA/WA-S3056/2020\|EPI_ISL_806986\|2020-08-21 | 2020-08 | Washington | WT* | WT | N501T | D614G |
| hCoV-19/USA/WA-S3043/2020\|EPI_ISL_806973\|2020-08-20 | 2020-08 | Washington | WT | WT | N501T | D614G |
| hCoV-19/USA/WA-S3042/2020\|EPI_ISL_806972\|2020-08-20 | 2020-08 | Washington | WT | WT | N501T | D614G |
| hCoV-19/USA/WA-S2930/2020\|EPI_ISL_806860\|2020-08-26 | 2020-08 | Washington | WT | WT | N501T | D614G |
| hCoV-19/USA/NY-AECOM_113/2020\|EPI_ISL_826657\|2020-08-04 | 2020-08 | New York | WT | WT | N501T | D614G |
| hCoV-19/USA/MA-MGH-03101/2020\|EPI_ISL_765704\|2020-08-31 | 2020-08 | Massachusetts | WT | WT | N501T | D614G |
| hCoV-19/USA/MA-Broad_UMMS-00220/2020\|EPI_ISL_976938\|2020-08-18 | 2020-08 | Massachusetts | WT | WT | N501T | D614G |
| hCoV-19/USA/MA-Broad_UMMS-00176/2020\|EPI_ISL_976927\|2020-08-28 | 2020-08 | Massachusetts | WT | WT | N501T | D614G |
| hCoV-19/USA/MA-Broad_UMMS-00004/2020\|EPI_ISL_935524\|2020-08-31 | 2020-08 | Massachusetts | WT | WT | N501T | D614G |
| hCoV-19/USA/MA-Broad_UMMS-00003/2020\|EPI_ISL_935523\|2020-08-29 | 2020-08 | Massachusetts | WT | WT | N501T | D614G |
| hCoV-19/USA/NY-MSHSPSP-PV19363/2020\|EPI_ISL_802301\|2020-09-29 | 2020-09 | New York | WT | WT | N501T | D614G |
| hCoV-19/USA/NY-MSHSPSP-PV18763/2020\|EPI_ISL_802255\|2020-09-17 | 2020-09 | New York | WT | WT | N501T | D614G |
| hCoV-19/USA/NY-MSHSPSP-PV18395/2020\|EPI_ISL_801870\|2020-09-09 | 2020-09 | New York | WT | WT | N501T | D614G |
| hCoV-19/USA/NY-AECOM_117/2020\|EPI_ISL_826660\|2020-09-01 | 2020-09 | New York | WT | WT | N501T | D614G |
| hCoV-19/USA/MA-MASPHL-00715/2020\|EPI_ISL_692948\|2020-09-08 | 2020-09 | Massachusetts | WT | WT | N501T | D614G |
| hCoV-19/USA/MA-Broad_UMMS-00186/2020\|EPI_ISL_976931\|2020-09-17 | 2020-09 | Massachusetts | WT | WT | N501T | D614G |
| hCoV-19/USA/MA-Broad_UMMS-00160/2020\|EPI_ISL_976919\|2020-09-19 | 2020-09 | Massachusetts | WT | WT | N501T | D614G |
| hCoV-19/USA/MA-Broad_UMMS-00150/2020\|EPI_ISL_976913\|2020-09-13 | 2020-09 | Massachusetts | WT | WT | N501T | D614G |
| hCoV-19/USA/MA-Broad_UMMS-00005/2020\|EPI_ISL_935526\|2020-09-03 | 2020-09 | Massachusetts | WT | WT | N501T | D614G |
| hCoV-19/USA/CT-Yale-1286/2020\|EPI_ISL_1080462\|2020-09-03 | 2020-09 | Connecticut | WT | WT | N501T | D614G |
| hCoV-19/USA/CT-Yale-455/2020\|EPI_ISL_729735\|2020-09-12 | 2020-09 | Connecticut | WT | WT | N501T | D614G |
| hCoV-19/USA/CT-QDX-4500/2020\|EPI_ISL_937037\|2020-09-29 | 2020-09 | Connecticut | WT | WT | N501T | D614G |
| hCoV-19/USA/CT-QDX-4499/2020\|EPI_ISL_937036\|2020-09-15 | 2020-09 | Connecticut | WT | WT | N501T | D614G |
| hCoV-19/USA/WI-WSLH-200723/2020\|EPI_ISL_631479\|2020-10-03 | 2020-10 | Wisconsin | G142D | WT | N501T | D614G |
| hCoV-19/USA/WI-WSLH-200722/2020\|EPI_ISL_631478\|2020-10-03 | 2020-10 | Wisconsin | G142D | WT | N501T | D614G |
| hCoV-19/USA/WI-WSLH-200721/2020\|EPI_ISL_631477\|2020-10-03 | 2020-10 | Wisconsin | G142D | WT | N501T | D614G |
| hCoV-19/USA/UT-UPHL-2012365234/2020\|EPI_ISL_745648\|2020-10-21 | 2020-10 | Utah | G142D | WT | N501T | D614G |
| hCoV-19/USA/TX-HMH-MCoV-15117/2020\|EPI_ISL_786601\|2020-10-17 | 2020-10 | Texas | WT | WT | N501T | D614G |

| | | | | | | |
|---|---|---|---|---|---|---|
| hCoV-19/USA/NY-NYCPHL-001123/2020\|EPI_ISL_671706\|2020-10-22 | 2020-10 | New York | WT | WT | N501T | D614G |
| hCoV-19/USA/NY-NYCPHL-001121/2020\|EPI_ISL_671704\|2020-10-22 | 2020-10 | New York | WT | WT | N501T | D614G |
| hCoV-19/USA/NY-NYCPHL-001058/2020\|EPI_ISL_633065\|2020-10-09 | 2020-10 | New York | WT | WT | N501T | D614G |
| hCoV-19/USA/NY-NYCPHL-001053/2020\|EPI_ISL_633063\|2020-10-13 | 2020-10 | New York | WT | WT | N501T | D614G |
| hCoV-19/USA/NY-MSHSPSP-PV19680/2020\|EPI_ISL_802371\|2020-10-08 | 2020-10 | New York | WT | WT | N501T | D614G |
| hCoV-19/USA/MI-MDHHS-SC22140/2020\|EPI_ISL_614191\|2020-10-06 | 2020-10 | Michigan | G142D | F486L | N501T | D614G |
| hCoV-19/USA/MI-MDHHS-SC22125/2020\|EPI_ISL_614176\|2020-10-06 | 2020-10 | Michigan | WT/G142D | F486L | N501T | D614G |
| hCoV-19/USA/MA-MGH-03876/2020\|EPI_ISL_1011513\|2020-10-27 | 2020-10 | Massachusetts | G142 | WT | N501T | D614G |
| hCoV-19/USA/MA-MGH-02843/2020\|EPI_ISL_765660\|2020-10-26 | 2020-10 | Massachusetts | G142 | WT | N501T | D614G |
| hCoV-19/USA/MA-MASPHL-00914/2020\|EPI_ISL_692834\|2020-10-22 | 2020-10 | Massachusetts | G142 | WT | N501T | D614G |
| hCoV-19/USA/LA-EVTL1833/2020\|EPI_ISL_1039534\|2020-10-09 | 2020-10 | Louisiana | G142 | WT | N501T | D614G |
| hCoV-19/USA/CT-QDX-3402/2020\|EPI_ISL_884321\|2020-10-29 | 2020-10 | Connecticut | G142 | WT | N501T | D614G |
| hCoV-19/USA/CT-QDX-3400/2020\|EPI_ISL_884319\|2020-10-30 | 2020-10 | Connecticut | G142 | WT | N501T | D614G |

*WT: no mutation compared to NC_045512.2 Severe acute respiratory syndrome coronavirus 2 isolate Wuhan-Hu-1, complete genome
