## Supplementary Table 5 for "SARS-CoV-2 spike protein gene variants with N501T and G142D mutation dominated infections in minks in the US"

Supplementary Table 5. Incidence of N501Y and N501T mutations in US and Canadain SARS-CoV-2 sequences

| Location | N501Y | | N501T | | Total* |
|---|---|---|---|---|---|
| US | 4,097 | 2.4% | 1,339 | 0.77% | 173,277 |
| Canada | 315 | 1.60% | 12 | 0.06% | 19,529 |
| World | 162,378 | 21.30% | ND** | ND | 760,644 |

*Sequences were downloaded from GISAID on March 12, 2021

**Not determined
